# Redeployment and training of healthcare professionals to Intensive Care during COVID-19: a systematic review

**DOI:** 10.1101/2021.01.21.21250230

**Authors:** Norha Vera San Juan, Matthew Camilleri, John Paul Jeans, Alexandra Monkhouse, Georgia Chisnall, Cecilia Vindrola-Padros

## Abstract

**Background:** A rapid influx of patients to intensive care and infection control measures during the COVID-19 pandemic required the rapid development of innovative redeployment and training strategies.

**Methods:** We conducted a systematic search of 9 databases including key terms related to intensive care AND training AND redeployment AND healthcare workers. Analysis consisted of a narrative synthesis of quantitative study outputs, and a framework-based thematic analysis of qualitative study outputs and grey literature. These results were then combined applying an interpretative synthesis.

**Results:** Twenty papers were analysed. These took place primarily in the UK (N=8, 40%) and USA (N=5, 25%). Themes included in the results are Redeployment: Implementation strategies and learnings; Redeployed staff experience and strategies to address their needs; Redeployed staff learning needs; Training formats offered and training evaluations; and Future redeployment and training concerns. Some of the redeployment implementation and training strategies documented in this review are: Skills-based redeployment, buddy support systems, and agreeing on locally-specific principles, rather than strict procedures.

**Conclusion:** The COVID-19 pandemic presented unique challenges to deliver training promptly while following infection control recommendations and develop flexible redeployment strategies. This study synthesises original approaches to tackle these challenges which are relevant to inform the development of targeted and adaptative training and redeployment plans.

## Introduction

To accommodate for the rapid fluctuations in the number of COVID-19 patients, healthcare organisations have been forced to optimise resource and staff allocation procedures. The unprecedented increase in the demand for intensive care services due to the COVID-19 pandemic involved a rapid redeployment of healthcare staff to these units. This posed multiple challenges, including devising new ways of working and rapid development and delivery of training. This review aimed to document key aspects of redeployment implementation, staff experiences and training.

Redeployment of staff from other specialties to intensive care aims to achieve the sustainable delivery of patient care by providing adequate patient care while still maintaining staff wellbeing standards (Hettle et al., 2020; NHS England & NHS Improvement, 2020a). Redeployment can be used to facilitate the daily work of Intensive Care Units (ICU) when implementing task-based models, where key tasks of patient care (i.e. hygiene) are carried out promptly by competent redeployed staff (Doyle et al., 2020). Staff redeployment can also help to address staffing gaps caused by staff sickness and previous vacancies.

Building staff competence and confidence is an essential principle of safe redeployment (NHS England & NHS Improvement, 2020b). Induction training must be provided to reacquaint redeployed staff with ICU ward multidisciplinary team practice and introduce them to clinical practices to care for COVID-19 patients and adequate use of Personal Protective Equipment (PPE) (Faderani et al., 2020). Time constraints and infection control measures pose unprecedented logistical challenges for training delivery, with traditional training methods such as those imparted in classrooms or at conferences not being possible.

The impact of redeployment on staff wellbeing has also been identified. Redeployed staff have expressed concern in relation to their safety, the impact of their work on family members (including infecting them with the virus) and their own training and career progression (Salem et al., 2020; Vera San Juan et al., 2021).

A range of novel strategies to implement redeployment and share knowledge in the context of the pandemic have been proposed and trialled. Gaining a detailed understanding of what worked and the needs that remain unaddressed will facilitate the development of redeployment plans.

### Aim

The aim of this review was to provide a detailed understanding of the characteristics of redeployment to ICU and training provision during the first year of the COVID-19 pandemic. It sought to identify what worked in redeployment and training, and what are concerns going forward with redeployment planning.

Research questions guiding the review were:

- What were the main strategies developed to redeploy staff to ICU?
- What were the principles of redeployment?
- What were redeployed staff experiences and perceived training needs?
- How were these needs addressed?
- What worked for redeployment and training?

## Methods

This review followed the Preferred Reporting Items for Systematic Reviews and Meta-Analyses (PRISMA) statement (Moher, Liberati, Tetzlaff, & Altman, 2009) and a protocol was developed a priori. The protocol was published on the authors’ institutional website (see Supplementary Material 1) as it was not eligible for publication on PROSPERO.

### Search strategy and study selection

Nine electronic databases were searched in December 2020 (including peer-reviewed and grey literature): Medline, CINAHL, PsychINFO and MedRxiv, Web of Science, The Health Management Consortium database, Social Science Research Network, OpenGrey and TRIP. The search strategy consisted of key terms referring to intensive care AND training AND redeployment AND healthcare workers. The search strategy was simplified when necessary for grey literature databases. A complete search strategy is provided in Supplementary Material 2.

Search results were imported into Rayyan (Ouzzani, Hammady, Fedorowicz, & Elmagarmid, 2016) and deduplicated. Title and abstract screening was conducted independently by two researchers (CV and NVSJ) and discrepancies were resolved via discussion until consensus was reached. Full texts of articles deemed relevant for inclusion were then screened against full review eligibility criteria. The references of included full-text articles were reviewed to identify additional articles.

### Eligibility criteria

Studies and commentaries published in peer reviewed journals or official reports were included in this review if they were focused on redeployment to Intensive Care Units and related wards during COVID-19. Publication date was restricted from December 2019 to 8^th^ of December 2020 (the date the search was conducted). There were no restrictions on language.

Articles were excluded if the focus was on redeployment to other areas of care, other viral infection emergencies, or changes in healthcare activities such as shifting to remote working.

### Data extraction and risk of bias assessment

NVSJ extracted the information of the articles onto a data extraction form developed on Qualtrics using a pre-defined list of data to extract (see Supplementary Material 1). The sections of the list relevant to redeployment and training were created after a preliminary scan of the selected articles.

Study details such as setting, population and methodological characteristics were collected from all articles. The core findings collected from the studies included details of redeployment experiences and implementation strategies (research question 1), and training programmes offered (research question 2). Special attention was paid to extract information about lessons learned and concerns for the future.

We expected a heterogeneous group of studies using different questions and outcomes, therefore the AACODS (Authority, Accuracy, Coverage, Objectivity, Date, Significance) checklist (Tyndall, 2010) was applied to assess the veracity of the source, clarity of the methods, acknowledgement of bias, and the relevance of the contribution to the field.

### Data synthesis method

We conducted a narrative synthesis of the study characteristics and quantitative study outputs (Popay et al., 2006) and a framework-based thematic analysis of qualitative study outputs and grey literature (Braun & Clarke, 2006; Gale, Heath, Cameron, Rashid, & Redwood, 2013). Quantitative and qualitative results were combined using an interpretative synthesis to develop an understanding of how they related and answer the two research questions (Barnett-Page & Thomas, 2009).

## Results

### Study selection

The screening and selection process is presented below in Figure 1 according to the PRISMA guidelines.

**Figure 1.**
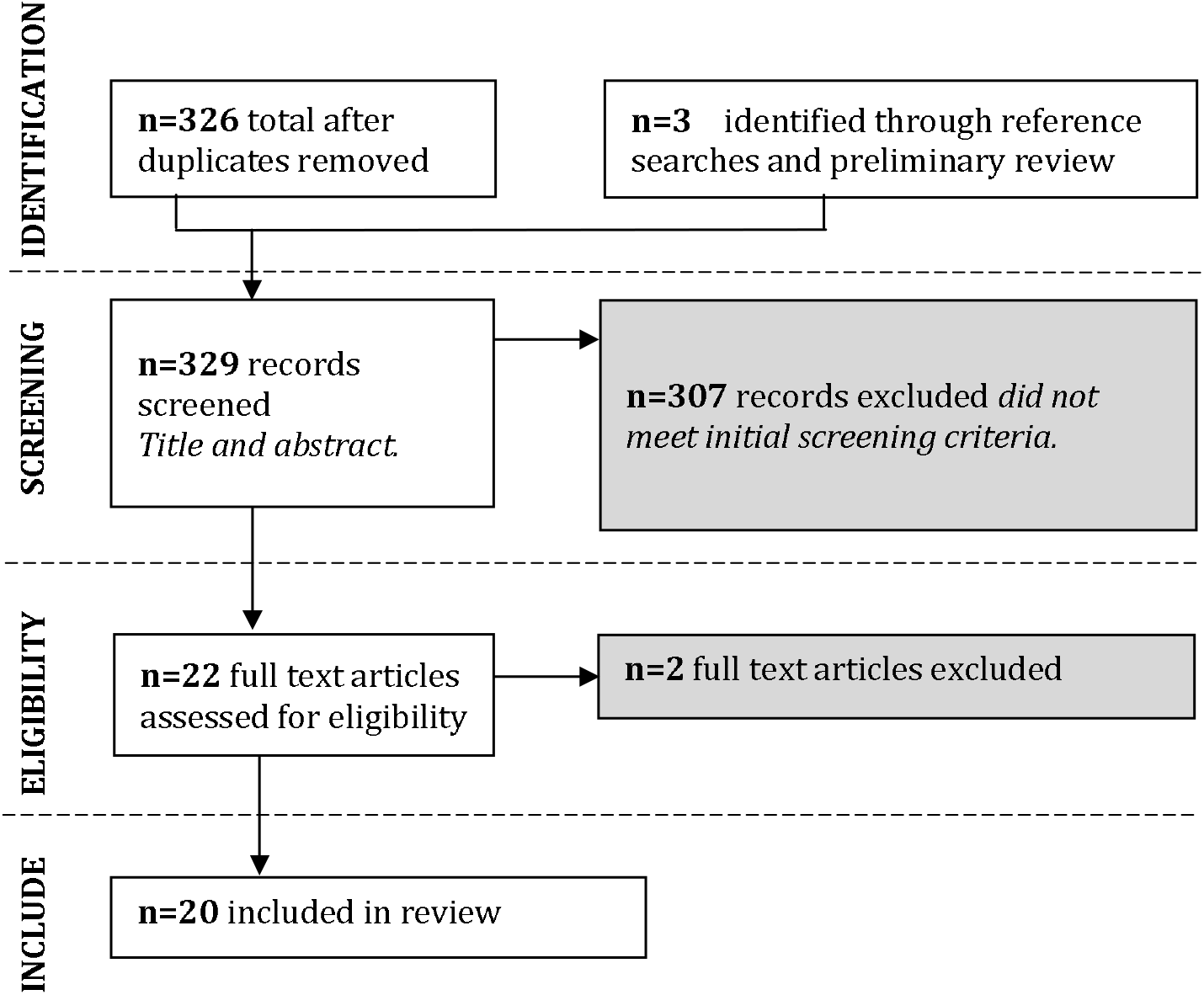
Preferred Reporting Items for Systematic Reviews and Meta-Analyses (PRISMA) flow diagram of the screening and selection process conducted in this systematic review.

Two articles were excluded at full text screening stage: Levy (2020), a commentary of one of the included articles which did not add information relevant to the research questions; and the study conducted by Chauhan (2020) which was not specific to redeployed staff.

### Study characteristics

From the twenty papers included in this systematic review, eleven (55%) were research studies; five (25%) were opinion pieces or commentaries; and four (20%) were reports or guidelines. Papers were primarily from the UK (N=8, 40%) and USA (N=5, 25%). Other locations included China, India, Germany and France. Eleven (55%) studies focused on redeployment implementation and experiences of redeployed staff, and nine (45%) on training delivery and evaluation, and dissemination of knowledge. The professional groups that were considered included nurses, anaesthetists, otolaryngologists, ophthalmologists, paramedics, orthopaedic surgeons, junior doctors, and physiotherapists.

From the eleven research studies included in the review, sample sizes ranged from ten (Marks, Edwards, & Jerge, 2020) to 1269 (Camilleri et al., 2020). One study reported 59% (N=19) of participants were male, and 41% (N=13) female (Payne, Rahman, Bullingham, Vamadeva, & Alfa-Wali, 2020). Other participant demographic characteristics such as ethnicity were not provided.

All studies met 80% or more of the quality criteria assessed. This is, they were written by recognised experts, included reference lists, targeted a clear aim and stated details such as date, location and limitations.

A full list of the studies included in this review can be found in Supplementary Material 3.

### Synthesis

Findings in this review are presented under the headings Redeployment:

Implementation strategies and learnings; Redeployed staff experience and strategies to address their needs; Redeployed staff learning needs; Training formats offered and training evaluations.

**Table 1** provides a summary of key findings.

**Table 1.**
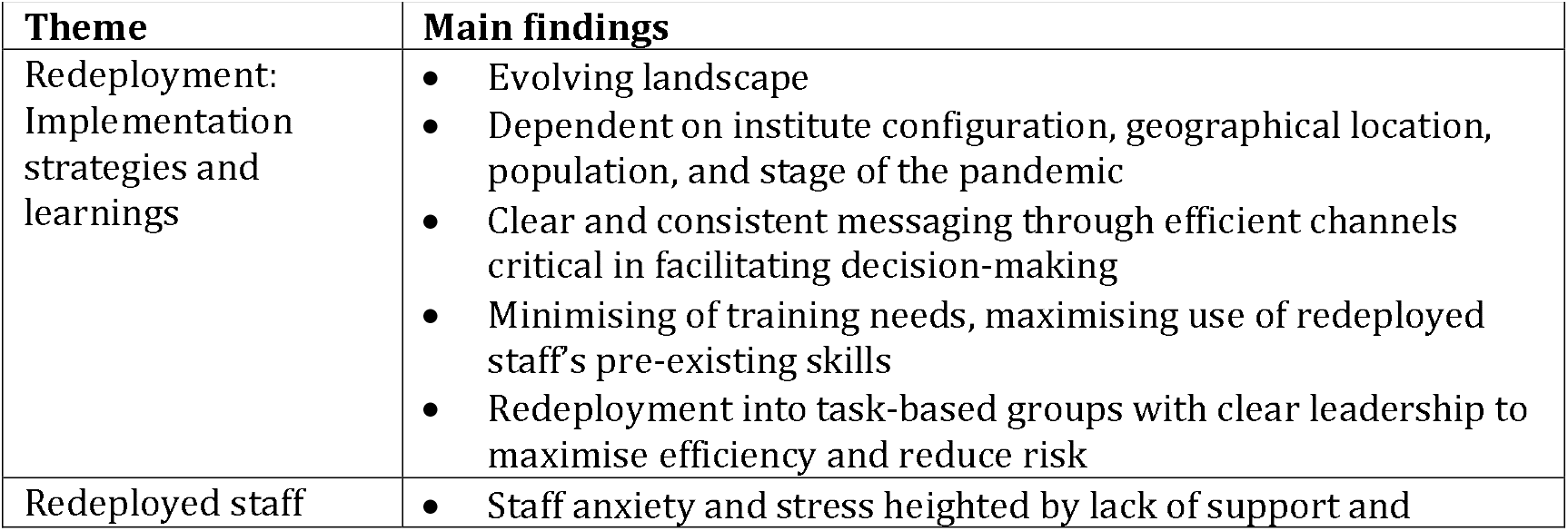
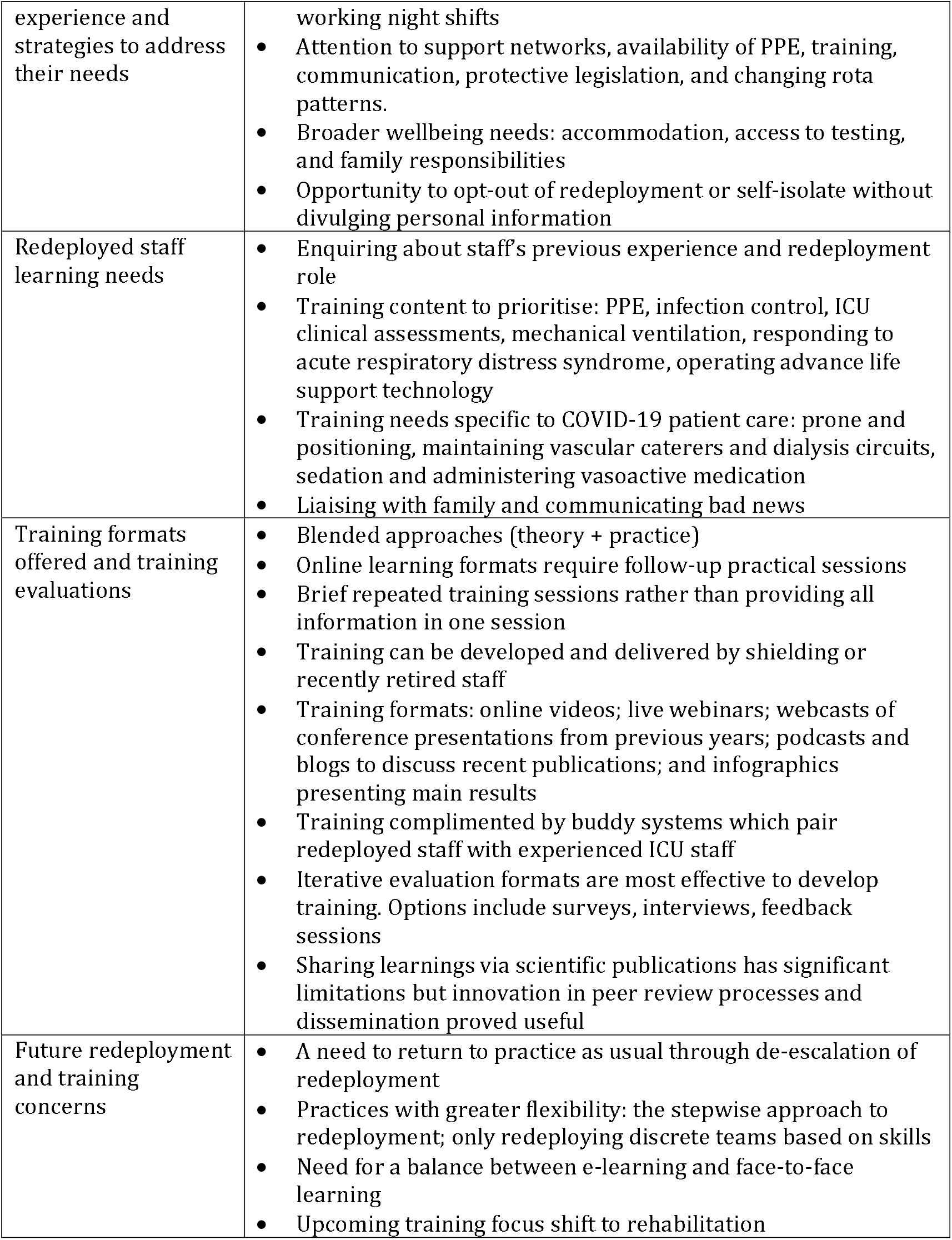
Summary of findings

#### Theme 1. Redeployment: Implementation strategies and learnings

By nature of the pandemic, areas of need were moving targets that constantly changed (Shipchandler, Nesemeier, Schmalbach, & Ting, 2020). Redeployment varied depending on the individual institution configuration, geographical and population context and stage along the pandemic (NHS England & NHS Improvement, 2020a).

Clear decision-making processes were facilitated, for example, by clear definitions of “urgent” and efficient channelling of information to create a clear and consistent message (George et al., 2020). For the latter, suggestions included limiting the use of email chains, using Dropbox or apps like Induction of Clinbee, and bespoke WhatsApp groups (NHS England & NHS Improvement, 2020a). In the UK, Regional Emergency Preparedness Response and Resilience (EPRR) teams were reported as key to assisting hospitals in the management of acute surge. Clinicians and managers should define warning triggers to anticipate a surge and establish communication lines with EPRR teams (NHS England & NHS Improvement, 2020a).

Redeployment planning aimed to minimise training needs and maximise use of redeployed staff’s previous knowledge by placing staff in roles where their existing skills could be more easily transferrable (Doyle et al., 2020). A successful approach to redeployment was allocating redeployed staff to task-based groups, that is, multidisciplinary teams with clear leadership and constant communication, that aimed to complete a specific necessary step of intensive care when requested by experienced ICU staff (Doussot et al., 2020; NHS England & NHS Improvement, 2020a). This represented an important shift in ways of working and understanding collaborations between health specialists (Doyle et al., 2020). There were benefits in some specialists taking over ICU roles, examples of this were Otolaryngologists examining epistaxis, peritonsillar abscess, and facial trauma (Shipchandler et al., 2020); experienced renal physicians, together with trainee Radiologists developing line-insertion teams, or Orthopaedists and Physiotherapists assisting with proning (Doyle et al., 2020). Benefits included reduction of personnel required for procedures, reduction of aerosolization of the virus, shorter time dedicated to procedures, and minimal or no training required for them to assist.

#### Theme 2. Redeployed staff experience and strategies to address their needs

Increasing staff buy-in was key for redeployment to work. Redeployed staff experienced anxiety and stress, particularly when lacking adequate support, during night shifts when less staff were available, and due to last minute rota changes. Staff wellbeing needs that required attention included accommodation, testing and family responsibilities (D’souza, Shetty, Apuri, & Moreira, 2020; George et al., 2020). Lim et al., (2020) reported that redeployed ophthalmologists’ anxiety reduced once their redeployment role began. This was attributed to receiving support from staff in the redeployed area, the sufficient availability of PPE, and adequate training.

Coughlan et al. (2020) described solutions for stressors that Junior Doctors experienced due to working in unfamiliar ICU settings. The interpersonal communication required for intense multidisciplinary teamwork was facilitated by visual aids, anonymised whiteboards and the use of walkie-talkies. Worries about potential negligence proceedings resulting from working beyond their usual competencies were mitigated by emergency legislation to protect doctors.

Guidelines developed by NHS England & NHS Improvement (2020a) proposed addressing redeployed staff’s needs by placing more experienced staff on nightshifts; encouraging questions; providing psychological support; accepting lower turnaround of patients; addressing issues about limited common areas and constant revision of rota patterns.

Finally, all staff should be given the opportunity to opt-out of redeployment and self-isolate without divulging any personal information (Burnett et al., 2020).

#### Theme 3. Redeployed staff learning needs

Redeployed staff learning needs varied depending on their previous experience and redeployment role. Training aimed to be targeted at the right level of difficulty depending on staff’s most recent work experience and focused solely on content that will be relevant for redeployed roles. An example of prioritising learning objectives was teaching non-specialists to recognize worsening conditions and the need for ventilation, while specialists mastered details of the operation of ventilators (Kuang et al., 2020).

Training needs considered essential to provide critical care services included learning basics of ICU monitoring such as conducting and interpreting systematic clinical assessments; mechanical ventilation; response to acute respiratory distress syndrome (i.e., intubation, and cardiac arrest); and operating advanced life support technology.

Additionally, staff providing care specifically to COVID-19 patients required an introduction to diagnosis and anticipated patient needs. These included prone and positioning, maintaining vascular catheters and dialysis circuits, sedation, and administering vasoactive medication (Camilleri et al., 2020; Doussot et al., 2020; NHS England & NHS Improvement, 2020b). Redeployed staff was also often asked to liaise with families and required training on communicating bad news (Payne et al., 2020).

A particular emphasis was made on learning needs related to PPE and infection control, especially during aerosol-generating procedures (D’souza et al., 2020; Doyle et al., 2020; Kuang et al., 2020).

#### Theme 4. Training formats offered and training evaluations

Most courses used blended approaches (theory + practice) and were collaboratively designed by a variety of clinical educators, intensive care experts and frontline staff (Camilleri et al., 2020; Hettle et al., 2020; Jansen et al., 2020; Riggall & Smith, 2015). Staff found it particularly useful when course content and practical sessions were repeated over time in consecutive sessions, rather than receiving an overwhelming amount of information in one session (Marks et al., 2020). This required allocating time to access sustained training and senior staff pointing to most up to date training available (Coughlan et al., 2020).

Training development and delivery could be helped forward by recruiting as facilitators staff who were shielding or had recently retired (Camilleri et al., 2020). Elder clinicians or those with other risk factors for severe COVID-19 infection could contribute to the pandemic response by leading simulation-based education sessions. Training formats included online videos; live webinars; webcasts of conference presentations from previous years; podcasts and blogs to discuss recent publications; and infographics presenting main results (Fawcett, Charlesworth, Cook, & Klein, 2020). Online learning formats allowed staff to access material at their own pace and check understanding, however, there was a need for practical follow-up sessions to consolidate learnings (Camilleri et al., 2020; Hettle et al., 2020; Marks et al., 2020).

In addition, courses were generally complemented by buddy systems (pairing up redeployed staff with more experienced ICU staff) or other similar set-ups to provide support for redeployed staff during practice (Doyle et al., 2020; Marks et al., 2020).

Most training programmes were evaluated through surveys or interviews and one article reported on the use of cycles of iteration composed of daily interactive feedback sessions with tutors and candidates to enable rapid improvement (Camilleri et al., 2020).

Sharing knowledge through scientific publications was thought to have significant limitations in the context of the pandemic. However, innovations in the peer review process and format of presenting results helped facilitate the continuation of this mode of learning (Fawcett et al., 2020). Despite these efforts, reaching a single message for best practice was a significant challenge during the pandemic and affected training development. Guidelines recommended teaching principles rather than strict procedures, as well as agreeing on locally-specific instructions (NHS England & NHS Improvement, 2020a).

#### Theme 5. Future redeployment and training concerns

As the pandemic evolved, the numbers of COVID-19 patients fluctuated rapidly and a need to return to practice as usual when possible has become more apparent. For this reason, redeployment implementation strategies have started to focus on facilitating de-escalating and escalating redeployment when necessary. Examples of practices which allowed for greater flexibility were the stepwise approach to redeployment, or only redeploying discrete teams based on skills (as described in the theme Redeployment: Implementation strategies and learnings) (Burnett et al., 2020; Doyle et al., 2020).

Regarding training, a need for balance between e-learning and face-to-face learning was identified. While e-learning is considered to have significant advantages such as a lower cost and reduced environmental impact, many feel there is still a need for face-to face learning. There are aspects of in-person meetings that enhance learning and wellbeing such as social interaction, hands-on teaching, and the opportunity to travel and visit venues, often incorporating some family downtime before or after the meeting (Fawcett et al., 2020).

Lastly, Camilleri et al. (2020) pointed out that as COVID-19 cases evolve, training focus will shift to rehabilitation.

## Discussion

This review synthesised data from twenty studies to identify the core aspects of redeployment implementation, redeployed staff experiences and training during the first year of the COVID-19 pandemic.

Key principles for successful redeployment were developing staff work groups based on skills rather than specialty; maximising use of redeployed staff’s transferable skills to minimise training; having a supportive environment; and developing flexible arrangements that allowed for scaling redeployment up or down. How best to implement these principles depended on each individual institution’s context, facilities, equipment, and the stage of the pandemic.

Redeployed staff stressed the importance of counting with continuous support from more experienced staff. Inductions and sustained training were key and should be targeted to the right level depending on staff’s previous experience. Central content included basics of ICU monitoring, response to acute respiratory distress syndrome, prone and positioning, and PPE donning and doffing. Staff assessed positively blended training with consecutive sessions.

Challenges faced by redeployed staff during the COVID-19 pandemic overlapped with previous studies assessing experiences of junior staff in Emergency Departments, or staff redeployed to disaster and war zones (see Table 2) (Brewer & Ryan-Wenger, 2009; Craven, 2017; Marion, Charlebois, & Kao, 2016). Theyyunni et al. (2013) analysed reflections from medical students after their emergency medicine rotation. The most common themes involved novice anxiety around critically ill patients and intubation procedures, miscommunication with other staff, and challenges stemming from the tension between textbook medicine and complex social situations. A challenge specific to the current pandemic was PPE use and everchanging guidelines. PPE use resulted in difficulties with communication, responding to emergencies in a timely manner, and increased physical burden (Scott & Unsworth, 2020). Other challenges were aspects outside of clinical practice that had an impact on redeployed staff’s wellbeing, such as, access to breakout rooms and PCR testing, and family caring responsibilities (Vera San Juan et al., 2021).

**Table 2.** Key challenges for redeployment and training during the COVID-19 pandemic

|  |
| --- |
| ▪ Efficient communication of clear and consistent messages in a rapidly changing environment.<br>▪ Challenges outside of clinical practice: family caring responsibilities, closure of leisure spaces, and more.<br>▪ PPE availability and use.<br>▪ Delivering training in a timely manner while following infection control recommendations.<br>▪ Establishing flexible redeployment strategies to adapt to unprecedented variations in the number of patients requiring intensive care. |

The essential role of training and need for innovative approaches has also been highlighted in the literature from previous virus outbreaks (Gertler et al., 2018). Multimodal, multidisciplinary and realistic simulation ran in consecutive sessions are recommended options for successful training (Darby, Whiting, & Winters, 2011; Ellington et al., 2020; Khamali et al., 2018; Riggall & Smith, 2015). Doulias, Gallo, Rubio-Perez, Breukink, and Hahnloser (2020) summarised changes in surgical training during the COVID-19 pandemic. Key aspects of successful training overlapped with those identified for redeployment to ICU. The necessary shift to e-learning was possible due to innovative and collaborative approaches that mitigated the loss of learning exposure during this time. In particular, interactive surgical simulation platforms offered a model of mentoring and continued guidance. Consultant bodies were more available to impart simulation training and engage in discussions with trainees due to the reduced elective surgery services.

Recommendations for future redeployment plans are moving towards flexible, innovative, and adaptive approaches. In the UK, the initial focus of the National Health Service (NHS) response to COVID-19 was establishing critical care capacity; this is now shifting towards developing pathways to support people to continue their rehabilitation and assessment in community settings (NHS England & NHS Improvement, 2020b; NHS England, NHS Improvement, & Health Education England, 2020). This will require staff receiving training on regular remote monitoring, communication with patients and family, and remote end of life care.

## Conclusion

Tackling ongoing challenges for healthcare provision in the current pandemic will require intense collaboration from multidisciplinary teams to build organisational resilience and optimise resources through successful execution of redeployment and training. Literature about healthcare provision in disaster contexts and warzones is the closest example of rapid redeployment to emergency care and can be a source of useful recommendations (such as the references cited in the discussion). However, the COVID-19 pandemic presented unique challenges and has resulted in original and innovative approaches which have been documented in this review.

## Data Availability

Search strategies and full citations of the papers included in this review are included in the supplementary materials.

