## Supplementary materials 1, 2, 3 for "Redeployment and training of healthcare professionals to Intensive Care during COVID-19: a systematic review"

Supplementary Material 1. Study Protocol

Review title.

The redeployment of healthcare professionals to intensive care units (ICUs) during COVID-19 and the implementation of training support

3. * Anticipated or actual start date.

15 September, 2020

4. * Anticipated completion date.

31 June 2021

Review question.

*State the review question(s) clearly and precisely. It may be appropriate to break very broad questions down into a series of related more specific questions. Questions may be framed or refined using PI(E)COS or similar where relevant.*

1. Redeployment
a. How was deployment/redeployment of staff defined during the COVID-19 pandemic?
b. At what levels were staff redeployed? (hospital, local, regional, national)
c. What were the main strategies developed to redeploy staff to ICUs?
2. Training
a. What were the main training needs of redeployed staff?
b. How did hospitals attempt to meet these needs?
c. Were any training programmes developed at regional levels?
d. Were any training programmes evaluated?

16. * Searches.

*State the sources that will be searched (e.g. Medline). Give the search dates, and any restrictions (e.g. language or publication date). Do NOT enter the full search strategy (it may be provided as a link or attachment below.)*

Scientific databases:

- MEDLINE
- CINAHL Plus
- PsychInfo
- MedRxiv

Grey literature databases:

- Web of Science
- Social Science Research Network (SSRN)
- The Health Management Information Consortium (HMIC) database
- TRIP
- OpenGrey

Authors of relevant conference presentations identified in these searches will be contacted to enquire about possible unpublished studies.

Publication date will be restricted from December 2019 – present time.

There will be no language restrictions

MedRxiv search will be limited to the first 5 pages of output.

Condition or domain being studied.

*Give a short description of the disease, condition or healthcare domain being studied in your systematic review.*

Redeployment of healthcare professionals to intensive care and emergency services during COVID-19

19. * Participants/population.

*Specify the participants or populations being studied in the review. The preferred format includes details of both inclusion and exclusion criteria.*

Inclusion

- Professionals redeployed to Intensive Care Units, A&E, Acute Medical Units and related wards during COVID-19.

Exclusion

- Healthcare professionals redeployed to other areas of care.
- Redeployment during other viral infection emergencies.

20. * Intervention(s), exposure(s).

*Give full and clear descriptions or definitions of the interventions or the exposures to be reviewed. The preferred format includes details of both inclusion and exclusion criteria.*

Inclusion

- Redeployment during COVID-19

Exclusion

- Redeployment during other viral infection emergencies
- Other changes in healthcare professional activities (e.g. shift to remote working)

21. * Comparator(s)/control.

*Where relevant, give details of the alternatives against which the intervention/exposure will be compared (e.g. another intervention or a non-exposed control group). The preferred format includes details of both inclusion and exclusion criteria.*

This review will not include a comparator/control group.

22. * Types of study to be included.

*Give details of the study designs (e.g. RCT) that are eligible for inclusion in the review. The preferred format includes both inclusion and exclusion criteria. If there are no restrictions on the types of study, this should be stated.*

Inclusion

- Studies and commentaries published in peer reviewed journals: no restrictions based on study design (quantitative, qualitative, case studies and RCTs)
- Official reports and guidelines.

Exclusion

- Conference abstracts
- Grey literature such as personal blogs, commentaries from relatives/carers, or news articles
- Information on non-institutional or non-NGO websites

Context.

*Give summary details of the setting or other relevant characteristics, which help define the inclusion or exclusion criteria.*

This review will focus on Intensive Care Units, A&E, Acute Medical Units and related wards during COVID-19.

24. * Main outcome(s).

*Give the pre-specified main (most important) outcomes of the review, including details of how the outcome is defined and measured and when these measurements are made, if these are part of the review inclusion criteria.*

This review will provide a detailed understanding of the characteristics of redeployment of healthcare professionals during COVID-19. Potential outcomes will include:

- Principles and key objectives of deployment/redeployment of healthcare staff during COVID-19 (i.e. NHS guidelines for redeployment)
- Measures of staff experience and satisfaction with redeployment
- Results from pre/post tests in relation to training
- Evaluation of training programmes (i.e. Kirkpatrick model).

* Measures of effect

*Please specify the effect measure(s) for you main outcome(s) e.g. relative risks, odds ratios, risk difference, and/or 'number needed to treat.*

Not Applicable

25. * Additional outcome(s).

*List the pre-specified additional outcomes of the review, with a similar level of detail to that required for main outcomes. Where there are no additional outcomes please state ‘None’ or ‘Not applicable’ as appropriate* 
*to the review*

Not applicable

26. * Data extraction (selection and coding).

*Describe how studies will be selected for inclusion. State what data will be extracted or obtained. State how this will be done and recorded.*

Search results will be deduplicated and imported into Rayyan to manage the references for the tittle and abstract stages of the screening. Two researchers will review articles independently and discuss any discrepancies until consensus is reached.

At least two review authors will extract relevant information from included studies onto a data extraction form developed on REDCap using a pre-defined list of data to extract (see below). We will resolve any disagreement by discussion with a third member of the team.

The list of data to extract will be created after an initial review of the selected articles. We will extract descriptive data and any primary qualitative data or themes identified relating to redeployment and training for redeployed healthcare staff working in emergency services during COVID-19

Preliminary list of data to extract:

- Citation details
- Location of study
- Data collection methods / source of data (for secondary data analysis studies)
- Data analysis methods
- Population:
- Professional group
- Sample size
- Sociodemographic and socioeconomic characteristics of the sample (gender, age, educational level, income level, and ethnicity)
- For research question 1: redeployment
- Redeployment process
- Objectives
- Level at which it occurred (hospital, regional, national…)
- Implementation strategies
- Learnings – what worked
- Redeployed staff
- Training needs
- Obligations
- For research question 2: Training
- Training programmes offered/developed
- Training programme evaluations
- Learnings – what worked

27. * Risk of bias (quality) assessment.

*State which characteristics of the studies will be assessed and/or any formal risk of bias/quality assessment tools that will be used.*

We expect a heterogeneous group of studies using different questions and outcomes, therefore a Mixed Methods Appraisal Tool to assess if the research question, methods, results and discussion align, alongside quality criteria specific to quantitative, qualitative and RCT studies will be applied. We will apply the AACODS checklist (Authority, Accuracy, Coverage, Objectivity, Date, Significance) to evaluate grey literature.

28. * Strategy for data synthesis.

*Describe the methods you plan to use to synthesise data. This* ***must not be generic text*** *but should be* ***specific to your review*** *and describe how the proposed approach will be applied to your data. If meta- analysis is planned, describe the models to be used, methods to explore statistical heterogeneity, and software package to be used.*

We will synthesise qualitative study outputs applying framework-based thematic analysis (Braun & Clarke 2006; Gale et al. 2013), narrative synthesis of quantitative outputs (Popay et al. 2006), and an interpretative synthesis combining both results (Barnett-Page and Thomas, 2009).

29. * Analysis of subgroups or subsets.

*State any planned investigation of ‘subgroups’. Be clear and specific about which type of study or participant will be included in each group or covariate investigated. State the planned analytic approach.*

We do not anticipate it will be possible to conduct subgroup analysis. As part of our synthesis of results we will list differences between professional groups and locations.

Dissemination plans.

*Do you intend to publish the review on completion?*

Peer reviewed publication 
Blog article

Supplementary Material 2. Search strategy

**Ovid: Medline, Psychinfo, Embase, and HMIC:**

1: "Intensive care" OR "acute medical units" OR "critical care" OR "critically ill" OR "critical illness" OR "intensive treatment unit" OR "intensive therapy unit" OR "high dependency unit" or ICU or ITU or HDU

2. Training OR education OR course OR prepar* OR "clinical competence" OR "education intervention" OR "training intervention" OR Skill OR "competency-based education" OR "simulation" OR "simulation training"

3. Redeploy* OR deploy* OR reallocation OR reallocate

4. staff OR "Healthcare worker" OR HCW OR "healthcare professional" OR clinician OR nurse OR anaesthetist OR doctor OR trainee OR AHP OR allied health professional

Search 1 AND 2 AND 3 AND 4

Time limit Dec 2019-Dec 2020

**MedRxiv**

Redeploy* AND Covid-19

**Web of Science**

1: "Intensive care" OR "acute medical units" OR "critical care" OR "critically ill" OR "critical illness" OR "intensive treatment unit" OR "intensive therapy unit" OR "high dependency unit" or ICU or ITU or HDU

2. Training OR education OR course OR prepar* OR "clinical competence" OR "education intervention" OR "training intervention" OR Skill OR "competency-based education" OR "simulation" OR "simulation training"

3. Redeploy* OR deploy* OR reallocation OR reallocate

4. staff OR "Healthcare worker" OR HCW OR "healthcare professional" OR clinician OR nurse OR anaesthetist OR doctor OR trainee OR AHP OR allied health professional

Search 1 AND 2 AND 3 AND 4

Time limit to “last five years”

**OpenGrey and SSRN**

Redeploy* AND Covid-19

"coronavirus"

**TRIP**

"intensive care" AND training AND redeploy*

Supplementary Material 3. Full list of included studies

| **Full citation** | **Type of entry** | **Location** |
| --- | --- | --- |
| NHS Clinical guide for the management of surge during the coronavirus pandemic: rapid learning version 2  (2020). United Kingdom. Retrieved from https://www.england.nhs.uk/coronavirus/wp-content/uploads/sites/52/2020/03/C0167-specialty-guide-surge-based-on-current-hospital-experience-v2.pdf | Report / Guideline | United Kingdom |
| Camilleri, M., Zhang, X., Norris, M., Monkhouse, A., Harvey, A., Wiseman, A., ... & Sinmayee, S. (2020). Covid-19 ICU remote-learning course (CIRLC): Rapid ICU remote training for frontline health professionals during the COVID-19 pandemic in the UK. Journal of the Intensive Care Society, 1751143720972630. | Scientific paper | UK and Ireland |
| George, I., Salna, M., Kobsa, S., Deroo, S., Kriegel, J., Blitzer, D., ... & Takeda, K. (2020). The rapid transformation of cardiac surgery practice in the coronavirus disease 2019 (COVID-19) pandemic: insights and clinical strategies from a centre at the epicentre. European Journal of Cardio-Thoracic Surgery, 58(4), 667-675. | Scientific paper | USA |
| Doussot, A., Ciceron, F., Cerutti, E., du Mont, L. S., Thines, L., Capellier, G., ... & Brunel, A. S. (2020). Prone positioning for severe acute respiratory distress syndrome in COVID-19 patients by a dedicated team: a safe and pragmatic reallocation of medical and surgical work force in response to the outbreak. Annals of Surgery, 272(6), e311. | Scientific paper | France |
| Doyle, J., Smith, E. M., Gough, C. J., Haq, A., Willis, C., Stevenson, T., & Reljic, M. (2020). Mobilising a workforce to combat COVID-19: An account, reflections, and lessons learned. Journal of the Intensive Care Society, 1751143720971540. | Opinion piece / commentary | England |
| Jansen, G., Latka, E., Behrens, F., Zeiser, S., Scholz, S., Janus, S., ... & Borgstedt, R. (2020). Hospital paramedic. An interprofessional blended learning concept to qualify paramedics and medical personnel for deployment in intensive care units and emergency departments during the COVID-19 pandemic. Der Anaesthesist. | Scientific paper | Germany |
| Nair, S. S., & Kaufman, B. (2020). Simulation-Based Up-Training in Response to the COVID-19 Pandemic. Simulation in Healthcare, 15(6), 447-448. | Report / Guideline | USA |
| Lim, C., De Silva, I., Moussa, G., Islam, T., Osman, L., Malick, H., ... & Burgula, S. (2020). Redeployment of ophthalmologists in the United Kingdom during the coronavirus disease pandemic. European journal of ophthalmology. | Scientific paper | UK |
| Shipchandler, T. Z., Nesemeier, B. R., Schmalbach, C. E., & Ting, J. Y. (2020). Otolaryngologists’ role in redeployment during the COVID-19 pandemic: a commentary. Otolaryngology–Head and Neck Surgery, 0194599820926982. | Opinion piece / commentary | USA |
| Fawcett, W. J., Charlesworth, M., Cook, T. M., & Klein, A. A. (2020). Education and scientific dissemination during the COVID‐19 pandemic. | Opinion piece / commentary | UK |
| Marks, S., Edwards, S., & Jerge, E. H. (2020). Rapid deployment of critical care nurse education during the COVID-19 pandemic. Nurse Leader. | Scientific paper | USA |
| Kuang, M., Wu, J., Luo, Y., Xiao, H., Liang, R., Hu, W., ... & Xiao, H. (2020). Training and reployment of non-specialists is an effective solution for the shortage of health care workers in the COVID-19 pandemic. medRxiv. | Scientific paper | China, Wuhan |
| Burnett, G. W., Katz, D., Park, C. H., Hyman, J. B., Dickstein, E., Levin, M. A., ... & Hamburger, J. (2020). Managing COVID-19 from the epicenter: adaptations and suggestions based on experience. Journal of anesthesia, 1-8. | Opinion piece / commentary | USA |
| D’souza, B., Shetty, A., Apuri, N., & Moreira, J. P. (2020). Adapting a secondary hospital into a makeshift COVID-19 hospital: A strategic roadmap to the impending crisis. International Journal of Healthcare Management, 1-6. | Report / Guideline | India |
| Brickman, D., Greenway, A., Sobocinski, K., Thai, H., Turick, A., Xuereb, K., ... & Liu, S. I. (2020). Rapid Critical Care Training of Nurses in the Surge Response to the Coronavirus Pandemic. American Journal of Critical Care, 29(5), e104-e107. | Scientific paper | USA |
| Naik, N., Finkelstein, R. A., Howell, J., Rajwani, K., & Ching, K. (2020). Telesimulation for COVID-19 Ventilator Management Training With Social-Distancing Restrictions During the Coronavirus Pandemic. Simulation & Gaming, 1046878120926561. | Scientific paper | USA |
| Payne, A., Rahman, R., Bullingham, R., Vamadeva, S., & Alfa-Wali, M. (2020). Redeployment of surgical trainees to intensive care during the COVID-19 pandemic: evaluation of the impact on training and wellbeing. Journal of surgical education. | Scientific paper | London |
| COVID-19: lessons for junior doctors redeployed to critical care | Opinion piece / commentary | London |
| NHS England, & NHS Improvement. (2020). COVID-19: Deploying our people safely. Retrieved January 9, 2021, from www.england.nhs.uk/coronavirus/workforce | Report / Guideline | UK |
| Hettle, D., Sutherland, K., Miles, E., Allanby, L., Bakewell, Z., Davies, D., Dhonye, Y., et al. (2020). Cross-skilling training to support medical redeployment in the COVID-19 pandemic. Future Healthcare Journal, 7(3), e41. Royal College of Physicians. Retrieved January 9, 2021, from https://www.ncbi.nlm.nih.gov/pmc/articles/PMC7571763/ | Scientific paper | Bristol |
